# Confounder control in biomedicine necessitates conceptual considerations beyond statistical evaluations

**DOI:** 10.1101/2024.02.02.24302198

**Authors:** Vera Komeyer, Simon B. Eickhoff, Christian Grefkes, Kaustubh R. Patil, Federico Raimondo

## Abstract

Machine learning (ML) models hold promise in precision medicine by enabling personalized predictions based on high-dimensional biomedical data. Yet, transitioning models from prototyping to clinical applications poses challenges, with confounders being a significant hurdle by undermining the reliability, generalizability, and interpretability of ML models. Using hand grip strength (HGS) prediction from neuroimaging data from the UK Biobank as a case study, we demonstrate that confounder adjustment can have a greater impact on model performance than changes in features or algorithms. An ubiquitous and necessary approach to confounding is by statistical means. However, a pure statistical viewpoint overlooks the biomedical relevance of candidate confounders, i.e. their biological link and conceptual similarity to actual variables of interest. Problematically, this can lead to biomedically not-meaningful confounder-adjustment, which limits the usefulness of resulting models, both in terms of biological insights and clinical applicability. To address this, we propose a two-dimensional framework, the *Confound Continuum*, that combines both statistical association and biomedical relevance, i.e. conceptual similarity, of a candidate confounder. The evaluation of conceptual similarity assesses on a continuum how much two variables overlap in their biological meaning, ranging from negligible links to expressing the same underlying biology. It thereby acknowledges the gradual nature of the biological link between candidate confounders and a predictive task. Our framework aims to create awareness for the imperative need to complement statistical confounder considerations with biomedical, conceptual domain knowledge (without going into causal considerations) and thereby offers a means to arrive at meaningful and informed confounder decisions. The position of a candidate confoudner in the two-dimensional grid of the *Confound Continuum* can support informed and context-specific confounder decisions and thereby not only enhance biomedical validity of predictions but also support translation of predictive models into clinical practice.

## 1. The critical role of confounder control in biomedical research and machine learning

### 1.1 Limitations of conventional confounder adjustment in scientific research

Confounding effects present significant challenges in scientific research, as they can obscure the true relationships between independent (predictor) and dependent (outcome) variables, leading to biased results and erroneous conclusions. Confounders are broadly defined as variables that correlate with both the predictor and the outcome, yet are not of primary interest^1,2^, potentially creating spurious associations or masking real effects (see e.g.^2–8^ for in-depth technical elaborations). Many disciplines adopt a conventional approach to adjust for a standard set of confounders. For instance, in imaging genetics, researchers commonly control for confounders such as age, sex (including squared or interaction terms), genetic ancestry/population stratification, SNP-derived confounders, scanner sequences, and further imaging-related variables^9–13^. A similar practice exists in fields like psychology, where adjustments are often made for age, sex, socioeconomic status, and education. Classical examples of confounders in neuroimaging research include measurement artifacts^7,14–16^, site effects^17^, demographics^18–20^, or lifestyle factors^21^. Such typically considered default confounders are frequently included without thorough investigation or justification^22–26^. While these standardized adjustments aim to mitigate confounding, relying solely on convention can be problematic for several reasons. Inadequate removal of confounders can inflate effect sizes, as results may be driven by confounding signals rather than the true variable of interest^12^. Conversely, over-adjusting for too many variables can eliminate relevant signals and lead to unstable results^17,27,28^. Adjusting for variables that are actually a consequence of the predictor(s) (i.e. not real confounders) may even induce false associations that can mislead subsequent interpretation of results (Berkson’s paradox)^12,29–31^. Overall, relying on generic or context-agnostic confounder adjustments can result in suboptimal analyses^2,32^. A more refined approach to confounder selection and deconfounding, tailored to the specific goals of the research, is therefore essential^2,12,32^.

### 1.2 Confounding in machine learning

Machine Learning (ML) workflows are increasingly employed in both, biomedical research and applications and hold promise for personalized medicine. Based on large, high-dimensional and oftentimes multimodal data ML predictive models can aid the identification of biomarkers of health and disease and can support diagnosis, prognosis and treatment choice, targeted to individuals^33–35^. Successful examples are deep learning-based cancer diagnosis, subtyping and staging^36^, inflammatory disease risk prediction^37^ or neuroimaging based predictive models outperforming DSM/ICD-based diagnoses^38^. However, translation of promising models to real-world clinical applications still remains challenging, sometimes referred to as AI chasm^39–42^. The AI chasm stems from unreliable predictions^22,43–45^, challenges with reproducibility and replicability, non-interpretability^40^, and limited generalizability^46^ of models (for further challenges see e.g.^22,35,39,47,48^).

Confounding effects contribute significantly to these concerns through misleading predictions and interpretations^12,32,49^. Confounders can influence predictions in many ways. For example, in a neuroimaging context, a model predicting hand grip strength (HGS) from neuroimaging derived features could be primarily driven by sex, i.e. men on average being stronger than women. Established tools to control for confounders at the level of study design, such as randomized control trials, restriction or matching^3^, may not be feasible in observational data as often used in ML workflows due to their advantageous sample size^2,38,49,50^. Consequently, post-hoc statistical approaches, such as (linear) confounder regression are commonly applied^4,7,12,32,51–53^. Alternatively, the contribution of confounders is often quantified by including them as predictors^7,12,54^.

In this paper, we highlight the pivotal role of confounder decisions in predictive modelling, showing that they can have a greater impact on model performance than feature selection or algorithm choice. While confounding is often treated as a purely statistical issue, we emphasize the need to align confounder decisions with the specific objectives of a study. This requires leveraging domain knowledge to assess the biological relevance and role of potential confounders. We propose a two-dimensional framework that combines statistical associations with biomedical conceptual relevance for evaluating candidate confounders. Although a comprehensive understanding of confounding effects ultimately requires a causal approach^55–57^, our goal here is to raise awareness of how integrating statistical and domain-specific insights can significantly improve the generalizability and scientific validity of neurobiomedical predictive models.

## 2. Confounder decisions can have a higher impact on model performance than feature or algorithm choice

To illustrate the importance of confounder decisions in predictive workflows, we conducted an exemplary supervised prediction of hand grip strength (HGS) from parcellated cortical, subcortical and cerebellar grey matter volume (GMV) features in the UK Biobank^58^ (see supplements for methods). HGS is an ideal target variable for this demonstration. It is reliable^59,60^ and eliminates further complexities associated with latent target measures such as intelligence or executive functioning scores. Additionally, HGS is an essential and ubiquitously employed assessment in clinical settings^61^.

First, we trained a *vanilla* model, using the GMV features, a linear ridge regression and no confounder adjustment in a 5-fold cross validation scheme (N_train_=22 276). Predictions on a previously completely hold out sample (OOS, N_test_=5 570) with this *vanilla* model yielded a Pearson correlation between true and predicted HGS of r=.64 (**Figure 1** -“Vanilla”). Changing the neuroimaging derived features from GMV to functional connectivity (FC) but keeping the rest of the setup constant led to a slight drop in performance to r=.58 between true and OOS predicted HGS (**Figure 1** – Feature impact, N_train_=3 337, N_test_=835). Starting off from the *vanilla* setup but this time changing the learning algorithm from a linear ridge regression to a non-linear support vector regression (SVR) with a hyper parameter optimized RBF kernel (see methods in supplementary materials) resulted in the same performance drop as for the feature change, i.e. r=.58 (**Figure 1** – Algorithm impact, N_train_=22 276, N_test_=5 570). However, using the *vanilla* setup but correcting the GMV features for the confounding variables *sex* and *age* by linearly regressing them out, rendered HGS (linearly) unpredictable, i.e. true and predicted HGS did not correlate anymore (r=.08) (**Figure 1** – Confounder impact, N_train_=18 593, N_test_=4 649).

**Figure 1.**
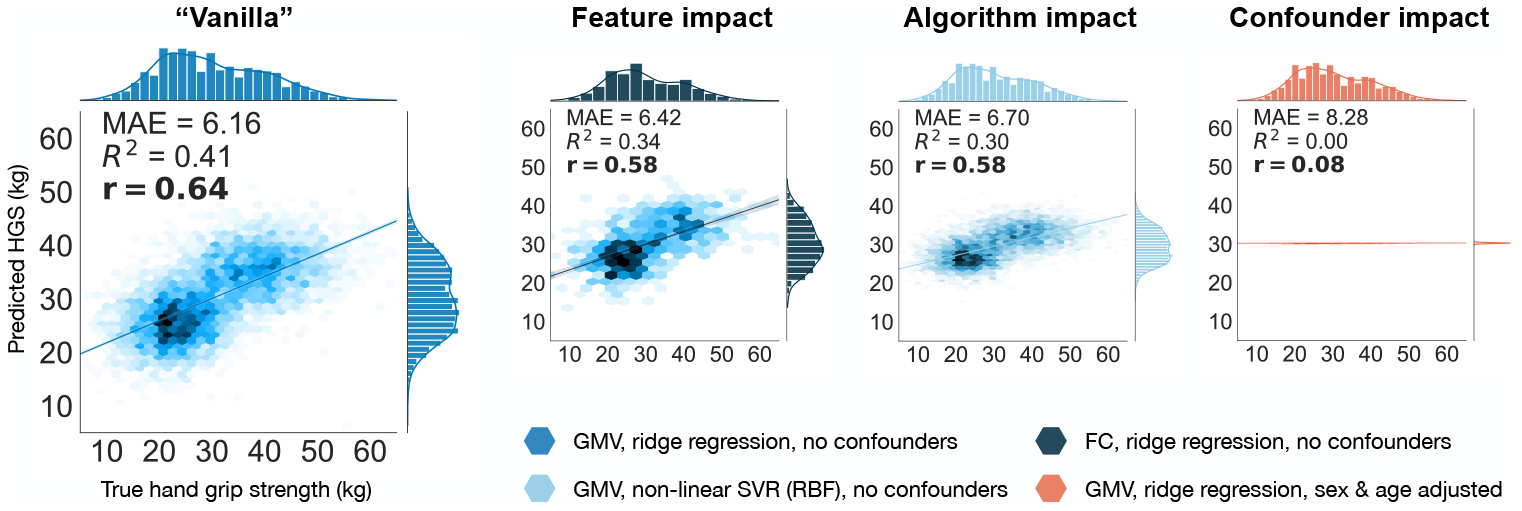
Impact of selection of neuroimaging derived feature, machine learning algorithm and confounder adjustment on out of sample (OOS) predictive performance. All plots show how accurate the respective model predicted the true, measured HGS. Performance is measured by Pearson correlation r between true and predicted HGS, coefficient of determination R^2^, and by mean absolute error (MAE). GMV: Gray matter volume, FC: Functional Connectivity, SVR: Support Vector regression, RBF: Radial Basis Function Kernel.

The choice of both, features and learning algorithm plays a crucial role in predictive modelling. Features should provide sufficient information about the target variable, and different learning algorithms can capture different aspects of the feature-target relationship, e.g. linear vs. non-linear relations, to different degrees. Therefore, tweaking the choice of features and learning algorithm in predictive workflows is important. Nonetheless, our illustrative analysis highlights the high impact of confounding variables on neurobiomedical predictive models. This renders informed decisions on confounder adjustment essential. Using the introduced predictive example, in the following we will elaborate on aspects to consider for informed confounder decisions.

## 3. A two-dimensional approach to confounding – an exemplary take

### 3.1. The statistical context of confound removal

Commonly, the relevance of a set of candidate confounders is determined by assessing their statistical association with the data. Variables with strong associations or high shared variance are considered as confounders in the predictive analysis. Performing statistical confounder evaluations is essential because one should only acknowledge confounding variables that share signal with the features and/or the target. This ensures that no confounder information is inadvertently introduced to the data by variables that do not share any signal^31^ and reveals variables which’s removal may enhance the signal to noise ratio of the feature-target relationship (high shared signal variables).

Following the traditionally established approach of seeing confounders primarily through the lense of association strengths, we exemplarily correlated 130 candidate confounding variables from the UKB with both HGS and GMV to evaluate their statistical association with the introduced GMV-HGS example (**Figure 2A**, Supplementary Figure S1B). We here illustrate the evaluation of statistical association strengths by means of correlations, but the particular statistical approach chosen can and should vary depending on the respectively wished insights (see e.g.^12^ for a broader approach). For illustration purposes in the following we mainly focus on a variable’s statistical association with the target. Focusing on a selection of example variables, body composition measures, sex and respiratory variables yielded highest absolute correlations with the target HGS. Variables such as “length of the working week in the main job” (LoWW), “systolic blood pressure”, “age” and “bone density” exhibited medium to small correlations with HGS (**Figure 2A**). Based on the absolute strength of their statistical association, candidate confounders can be ordered from low to high absolute association strength. This forms a statistical axis for confounder evaluations. For a set of example candidate confounders this could for instance be: scan-site < systolic blood pressure < age < LoWW ≈ bone density < sex.

**Figure 2.**
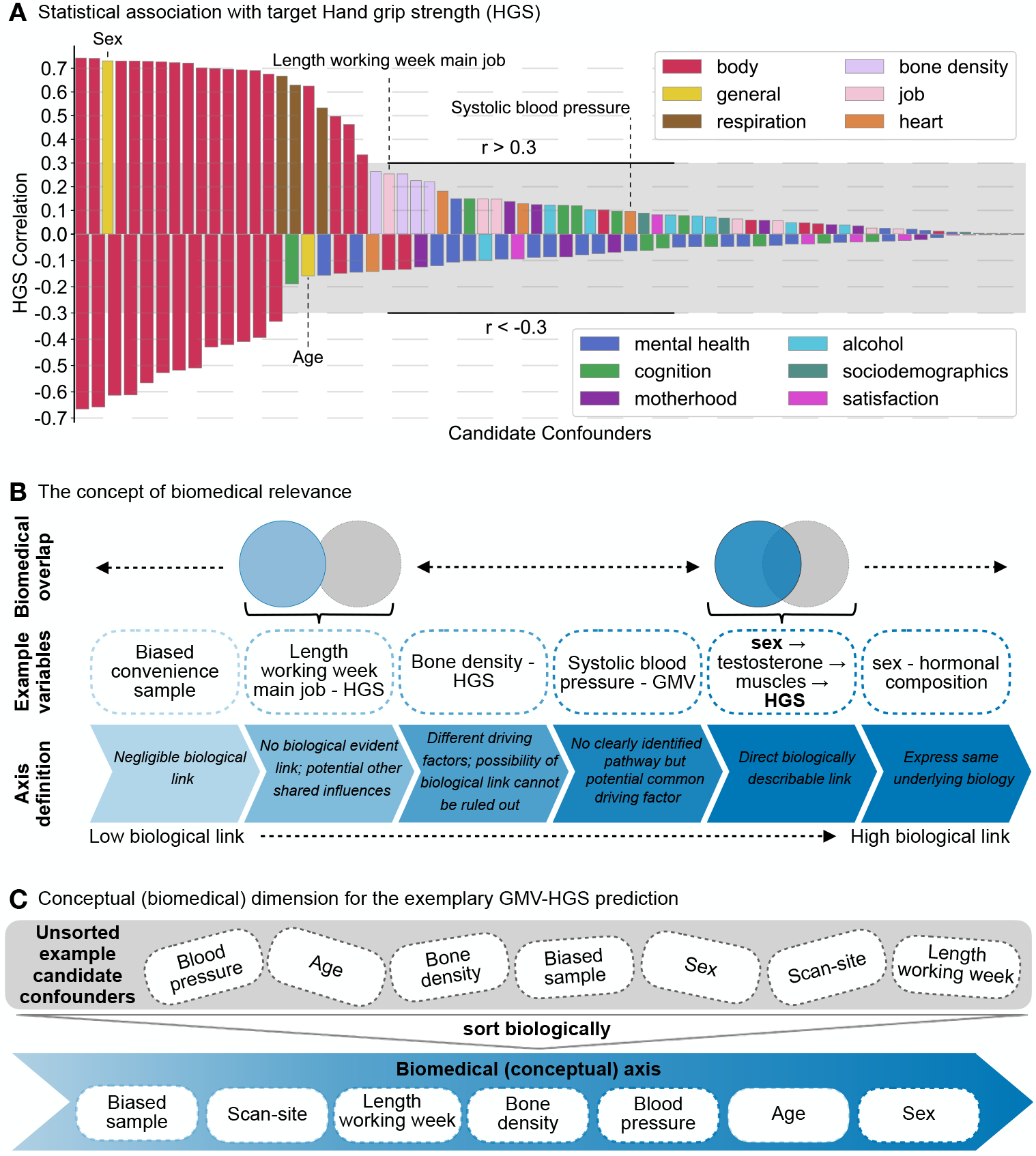
The principles of statistical and conceptual relevance of candidate confounding variables. **A)** Correlations of 130 summary behavioural variables that could potentially be considered as confounders with the exemplary target HGS. The variables were sorted into 12 higher-level categories. Correlations refer to Pearson’s r for continuous confounds, Spearman correlations for ordinal variables and point-biserial correlation coefficients for binary variables. Grey areas mark absolute correlations smaller than |r|=0.3 for illustration purposes. **B)** The general concept of biomedical relevance of two variables for each other. Two variables can share a different amount of biomedical information (top). The shared biology between sex and hand grip strength (HGS) for example is higher than between HGS and Length of working week (LoWW) (middle). The degree of biomedical overlap can be ranked on a conceptual axis, ranging from a negligible biological link to two variables expressing (almost) the same underlying biology (bottom). **C)** Seven exemplary candidate confounders for the GMV-HGS prediction task can be sorted according to their biological relevance for the prediction task, following the definition of the conceptual axis as specified in (B).

Having a broader look at all associations it becomes apparent that a huge variety of variables, in fact, almost all variables to some degree correlate with HGS. Especially in large datasets with many measured variables available, such as the UKB, almost everything is somewhat related to everything. Consequently, for meaningful evaluation, if a variable is relevant to be considered as a confounder, another dimension, independent of statistical considerations must be employed.

### 3.2. Consideration of potential biological relationships

Beyond the statistical association, however, variables can also be of different relevance for each other from a biomedical, i.e. conceptual point of view. This viewpoint relies on domain knowledge, expert experience and literature. Conceptually, two variables can express quasi the same biology or describe completely distinct biological phenomena. In extreme cases, one of the variables does not describe any biological phenomenon at all. For example, sex and hormonal composition describe almost the same underlying biology. Therefore, they strongly overlap in biomedical terms (**Figure 2B**). The same applies to sex and HGS, for which there is a directly describable biological link, namely sex → testosterone → muscle mass → HGS. On the other hand side, a measurement such as LoWW primarily does not describe a biological phenomenon and is therefore not directly biomedically meaningful for a second variable that exhibits biological meaning, such as HGS. In other words, LoWW and HGS may have shared influences, but per se do not overlap in their biological meaning. In the most extreme case, for instance with a biased convenience sample, there is no common biology whatsoever with any other biologically relevant variable as it is a purely technical influencing factor. Of course, using an appropriate data sample is highly relevant for any study, nonetheless this is not directly conceptually biologically linked to a variable that inherently carries biological meaning.

The varying degree of biomedical relevance (or overlap) of two variables for each other can be perceived as a continuum. This continuum can reach from a *negligible biological link* as the lowest extreme, to *expressing the same underlying biology* as the highest extreme. Thereby one can form an axis of increasing biomedical, conceptual linkage between two variables (**Figure 2B** bottom). The principal of biomedical relevance can be used to build a biomedical, conceptual axis for confounder evaluations. Candidate confounders can be evaluated and sorted on this axis based on their biological overlap with the feature and/or target variables, here with GMV and/or HGS (**Figure 2C**). If we take the same example set of candidate confounders as for the statistical illustration, the sorting in the biomedical, conceptual dimension could be: scan-site < LoWW < bone density < systolic blood pressure < age < sex.

Importantly, this clearly delineated suggested sorting serves as a simplified and illustrative example to explain the broader concept. However, in many practical applications, determining the exact order can be challenging. While extreme cases are often clear, the middle regions of the axis may be more ambiguous, leading to a final order that could appear somewhat subjective. For instance, in GMV-HGS predictions, factors such as sex and age clearly play a significant biological role. In contrast, the placement of variables such as bone density or blood pressure on the biomedical axis could vary, ultimately reflecting subjective judgment. This is not a major issue, as the precise ordering is unlikely to be critical. What is crucial, however, is that potential confounders are assessed based on their biomedical relevance to the specific scientific question, drawing on domain knowledge and a thorough review of the literature. The key point is the thinking about the biomedical meaning of a variable and its implications in a research endeavour - something that purely statistical analysis cannot capture. Incorporating domain expertise and formalizing it using the proposed conceptual axis enables the reasoning process to be traceable, i.e. the *why* behind including or excluding variables is made transparent.

### 3.3. The biomedical relevance of a confounder is independent of its statistical association strength

Both, statistical and biomedical evaluations of candidate confounders are essential as they evaluate complementary aspects of a candidate confounder’s relevance in a predictive task. Therefore, biomedical and statistical evaluations can be orthogonally combined in a two-dimensional (2D) grid. The statistical evaluation forms the vertical axis, while the biomedical conceptual evaluation builds the horizontal axis. Candidate confounders can be sorted in this 2D grid according to their statistical association strength and biomedical relevance as elaborated on in **3.1** and **3.2**, respectively. For the previously introduced example set of candidate confounders, the respective **statistical** ordering was *scan-site < systolic blood pressure < age < LoWW ≈ bone density < sex* and the **conceptual** ordering was *scan-site < LoWW < bone density < systolic blood pressure < age < sex*. Therefore, sex is positioned in the top right corner of the 2D grid, while scan-site is positioned in the bottom left corner. The other example candidate confounders can be sorted in this 2D grid following the same principle (**Figure 3A**). Sorting all previously statistically evaluated variables (**Figure 2A**) in this manner leads to a fully filled 2D *Confound Continuum* for the GMV-HGS prediction task (**Figure 3B)**.

**Figure 3.**
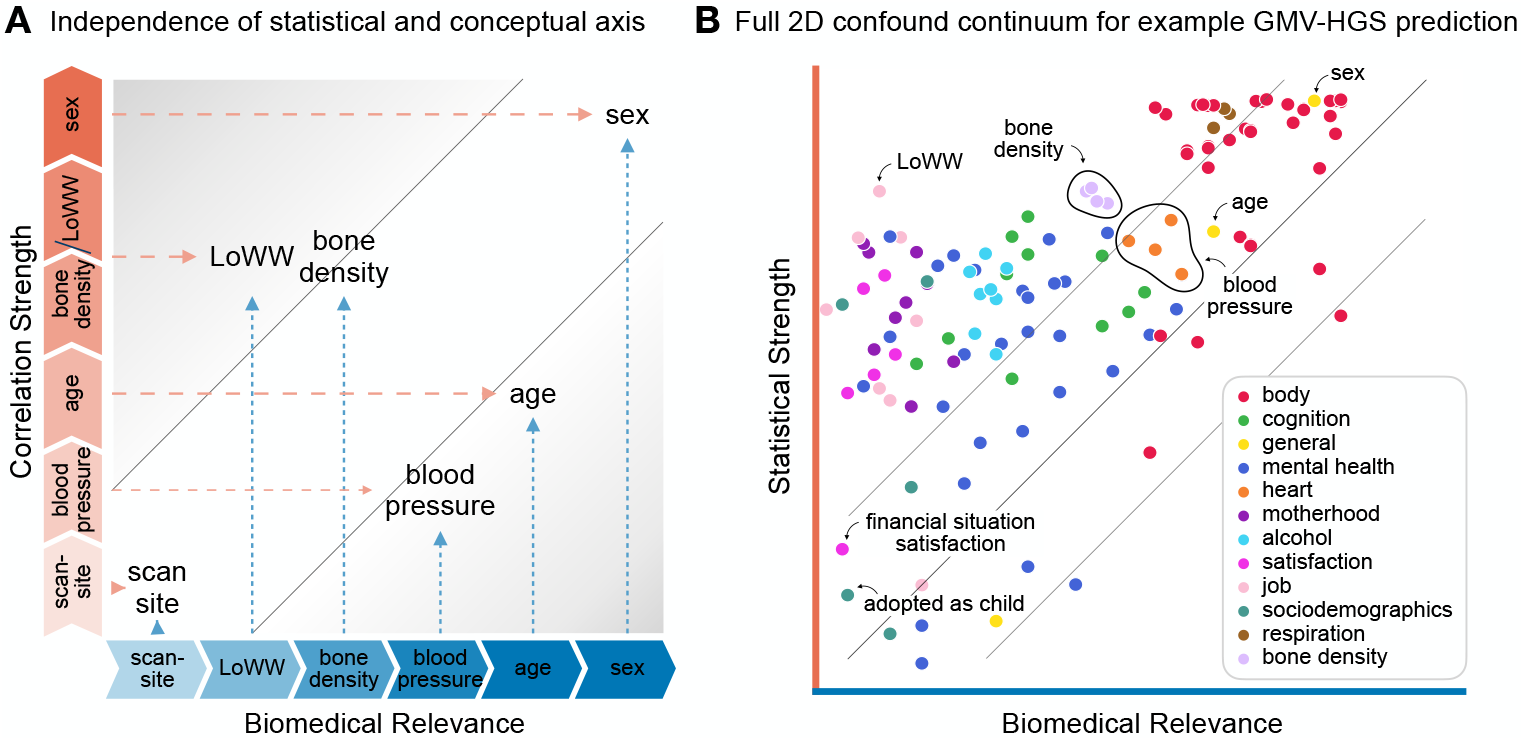
Two-dimensional Confound Continuum. **A)** Schematic of the 2D grid, combining the exemplary statistical and biomedical evaluations as performed in **3.1** and **3.2 (Figure 2)** as vertical and horizontal axis, respectively. Variables lying on the diagonal are similarly relevant from a statistical and conceptual perspective so that confounder decisions are clear. In contrast, variables in off-diagonal areas emphasize the independence of statistical and biomedical relevance. Such variables require to counterweigh the statistical and biomedical importance to make the most appropriate decision for a given prediction task. **B)** Full 2D Confound Continuum for the GMV-HGS prediction task taking into account all 130 variables introduced in the statistical evaluations **(3.1, Figure 2A)**.

While sex and scan-site lie on the diagonal in the example 2D space, the other example variables are positioned off-diagonal (**Figure 3A**). This emphasizes the independent and complementary nature of statistical and biomedical investigations. For instance, LoWW correlated with HGS with r=0.25. Following the ubiquitous definition of a confounder as variables that correlate with features and/or targets, but are not of primary interest, this statistical association could be considered strong enough to qualify LoWW as a confounder to adjust for. However, thinking about the biomedical meaning behind predicting HGS from GMV, the presumable goal would be to find out meaningful information in the brain that is predictive of an individual’s grip strength. With this biomedical goal in mind, correcting gray matter brain information for how long a person works per week does not appear to be biomedically meaningful. In contrast, age correlated with HGS only with |r|^1^=0.16, i.e. less than LoWW, and with GMV with |r|=0.20. However, when again keeping in mind the biomedical goal of a GMV-HGS prediction task, one might want to address age as a confounder because of its biological relevance for both HGS and GMV. To gain insights into gray matter brain information that is predictive for an individual’s grip strength, one therefore might want to make sure that indeed brain information without age influences was used for successful predictions. In other words, for variables on the diagonal the decision for or against adjustment is most often clear as it only depends on an individually set *importance* threshold, e.g. r=0.2. Variables in off-diagonal areas however alert to scenarios where it is crucial to counterweight the statistical and biomedical importance to make biomedically meaningful decisions on confounder adjustment.

In summary, statistical and biomedical conceptual evaluations of confounder influences are independent and must therefore both be taken into account. This can be facilitated by the introduced 2D *Confound Continuum*. Acknowledging that biomedical and statistical validity are distinct but complementary concepts can enhance the understanding of the role of confounders in a predictive task. The *Confound Continuum* thereby can facilitate and support informed decisions on confound removal through acknowledging problem-specific nuances.

## 4. Discussion

Confounders play a significant role in machine learning predictive workflows as they can have major impact on both performance and meaning of predictions without themselves being of primary interest. By exemplarily predicting hand grip strength (HGS) from gray matter volume (GMV) under varying conditions, we illustrated that confounder adjustments can have a greater impact on model performance than the choice of features or learning algorithm. Consequently, informed confounder decisions are indispensable to improve both interpretability and generalizability of machine learning models. Traditionally, confounders are often primarily approach through statistical association strengths. The here presented 2D framework aims to create awareness for the imperative need to complement such statistical considerations with biomedical, conceptual domain knowledge to arrive at meaningful and informed confounder decisions.

A key element of the suggested 2D approach, is addressing the semantic meaningfulness of confounder adjustment by assessing the biomedical validity of confound-adjusted features and targets as well as the biomedical meaning of resultant models and predictions. The here presented exemplary conceptual evaluations were tried to be kept clear example cases. However, candidate confounders may exhibit more complex networks of nested and cascadic conceptual relationships with the features and/or the target as well as with each other. Such cascadic influences can emerge because biomedical mechanisms usually form complex networks^24,66^. For instance, hormone levels (e.g. sex hormones) can influence body fat composition. However, body fat composition in conjunction with sex can also influence hormone levels^67^. Such interactions can affect the introduced cascade sex → testosterone → muscle growth → HGS. Additionally, body fat composition may further interact with respiratory capacities and thereby influence additional factors such as physical fitness. Consequently, even seemingly unrelated variables may indirectly impact the actual relationship between GMV and HGS. While pure statistical evaluations are blind to such cascadic mechanisms, the conceptual axis by design is meant to integrate exactly these types of interactions into confounder considerations.

Ultimately, the evaluation of biomedical relevance of a candidate confounder requires a causal analysis that investigates cause-effect relationships between involved variables, i.e. candidate confounding variables, features and the target. Such a causal analysis can result in a directed acyclic graph (DAG) or causal diagram that captures the network-like nature of biomedical mechanisms^24,56,57,66^. The here presented conceptual axis can be seen as a simplified version of such a causal investigation. Such a DAG allows to differentiate if there exists a path between two variables, the directionality of such a path, the connection strength of the two variables the path connects as well as the sum of all paths from one variable to another. In contrast, the here introduced conceptual dimension summarizes all these aspects in one axis. The biomedical conceptual axis thereby summarizes all causal paths and their respective causal strength, by combining the question about the existence of causal path and the corresponding direction and strength into the question of “how biomedically relevant is this candidate confounding variable?”. While this comes for the price of losing some details, it comes with the strength of allowing an expert-and experience-oriented intuitive approach. By introducing this biomedical conceptual axis we want to create awareness for the need of investigating the biomedical meaningfulness and role of candidate confounders additional and complementary to statistical evaluations and want to provide an intuitively usable framework that combines the statistical and conceptual perspective (for a more recipe-style tool leveraging DAGs see e.g. ^24,55^.

The *Confound Continuum* aims to support informed confound removal decisions in a problem-dependent manner, bridging the gap between statistical and biomedical conceptual perspectives. It emphasizes that biomedical and statistical validity are distinct concepts and connects confound removal to model interpretation. In the realm of biology, no variable exists in complete isolation from others. Certain datasets might create the impression of some variables being biologically unrelated, but this likely reflects the inherent limitations of any dataset, which can only capture a finite number of measured variables. Conversely, in wide-enough data, most measured variables will be multi-correlated from a statistical perspective and (cascadically) inter-related from a conceptual viewpoint. This multi-correlatedness can be exploited by methods such as meta-matching^68^, but renders pure statistical evaluations of confounders not sufficient. Therefore, it is crucial to dissect the interconnectedness of biological variables from a bio-conceptual perspective and combine this perspective with statistical data-insights to derive valid models and corresponding interpretations – a bridge provided by the *Confound Continuum*.

## Data Availability

All individual data used in this study were obtained from the UK Biobank, a major biomedical database (www.ukbiobank.ac.uk) under application number 41655, and are available to all approved UK Biobank researchers.

## 5. Acknowledgments

This research has been conducted using the UK Biobank Resource under application number 41655. This research was funded by the Deutsche Forschungsgemeinschaft (DFG, German Research Foundation) – Project-ID 431549029 - Collaborative Research Centre CRC1451 on motor performance project B05.

## Footnotes

1 Indication of absolute correlation value for comparability of association strengths.

## References

1. Pourhoseingholi MA, Baghestani AR, Vahedi M. How to control confounding effects by statistical analysis. Gastroenterol Hepatol Bed Bench. 2012;5(2):79–83.

2. Chyzhyk D, Varoquaux G, Milham M, Thirion B. How to remove or control confounds in predictive models, with applications to brain biomarkers. GigaScience. 2022;11:giac014. doi:10.1093/gigascience/giac014

3. Jager KJ, Zoccali C, MacLeod A, Dekker FW. Confounding: What it is and how to deal with it. Kidney Int. 2008;73(3):256–260. doi:10.1038/sj.ki.5002650

4. Kostro D, Abdulkadir A, Durr A, et al. Correction of inter-scanner and within-subject variance in structural MRI based automated diagnosing. NeuroImage. 2014;98:405–415. doi:10.1016/j.neuroimage.2014.04.057

5. MacKinnon DP, Krull JL, Lockwood CM. Equivalence of the Mediation, Confounding and Suppression Effect. Prev Sci. 2000;1(4):9.

6. McNamee R. Confounding and confounders. Occup Environ Med. 2003;60(3):227–234. doi:10.1136/oem.60.3.227

7. Rao A, Monteiro JM, Mourao-Miranda J. Predictive modelling using neuroimaging data in the presence of confounds. NeuroImage. 2017;150:23–49. doi:10.1016/j.neuroimage.2017.01.066

8. Zhao Q, Adeli E, Pohl KM. Training confounder-free deep learning models for medical applications. Nat Commun. 2020;11(1):6010. doi:10.1038/s41467-020-19784-9

9. Stein JL, Medland SE, Vasquez AA, et al. Identification of common variants associated with human hippocampal and intracranial volumes. Nat Genet. 2012;44(5):552–561.

10. McCaw ZR, Colthurst T, Yun T, et al. DeepNull models non-linear covariate effects to improve phenotypic prediction and association power. Nat Commun. 2022;13(1):241.

11. Duncan NW, Northoff G. Overview of potential procedural and participant-related confounds for neuroimaging of the resting state. J Psychiatry Neurosci. 2013;38(2):84–96.

12. Alfaro-Almagro F, McCarthy P, Afyouni S, et al. Confound modelling in UK Biobank brain imaging☆. Published online 2021:17.

13. Miller KL, Alfaro-Almagro F, Bangerter NK, et al. Multimodal population brain imaging in the UK Biobank prospective epidemiological study. Nat Neurosci. 2016;19(11):1523–1536. doi:10.1038/nn.4393

14. Geerligs L, Tsvetanov KA Cam-CAN, Henson RN. Challenges in measuring individual differences in functional connectivity using fMRI: The case of healthy aging: Measuring Individual Differences Using fMRI. Hum Brain Mapp. 2017;38(8):4125–4156. doi:10.1002/hbm.23653

15. Power JD, Barnes KA, Snyder AZ, Schlaggar BL, Petersen SE. Spurious but systematic correlations in functional connectivity MRI networks arise from subject motion. NeuroImage. 2012;59(3):2142–2154. doi:10.1016/j.neuroimage.2011.10.018

16. Satterthwaite TD, Wolf DH, Loughead J, et al. Impact of in-scanner head motion on multiple measures of functional connectivity: Relevance for studies of neurodevelopment in youth. NeuroImage. 2012;60(1):623–632. doi:10.1016/j.neuroimage.2011.12.063

17. Spisak T. Statistical quantification of confounding bias in predictive modelling. Published online November 1, 2021. Accessed January 31, 2023. http://arxiv.org/abs/2111.00814

18. Bugg JM, Zook NA, DeLosh EL, Davalos DB, Davis HP. Age differences in fluid intelligence: Contributions of general slowing and frontal decline. Brain Cogn. 2006;62(1):9–16. doi:10.1016/j.bandc.2006.02.006

19. Hartshorne JK, Germine LT. When Does Cognitive Functioning Peak? The Asynchronous Rise and Fall of Different Cognitive Abilities Across the Life Span. Psychol Sci. 2015;26(4):433–443. doi:10.1177/0956797614567339

20. Horn (1967) - age differences in fluid and crystallized intelligence.pdf.

21. Kahlert J, Gribsholt SB, Gammelager H, Dekkers OM, Luta G. Control of confounding in the analysis phase – an overview for clinicians. Clin Epidemiol. 2017;Volume 9:195–204. doi:10.2147/CLEP.S129886

22. Benkarim O, Paquola C, Park B yong, et al. The Cost of Untracked Diversity in Brain-Imaging Prediction. bioRxiv. Published online June 2021:34. doi:10.1101/2021.06.16.448764

23. Weinstein SM, Davatzikos C, Doshi J, Linn KA, Shinohara RT, For the Alzheimer’s Disease Neuroimaging Initiative. Penalized decomposition using residuals (PeDecURe) for feature extraction in the presence of nuisance variables. Biostatistics. Published online August 11, 2022:kxac031. doi:10.1093/biostatistics/kxac031

24. Wysocki AC, Lawson KM, Rhemtulla M. Statistical Control Requires Causal Justification. Advances in Methods and Practices in Psychological Science. 2022;5(2).

25. Becker TE. Potential Problems in the Statistical Control of Variables in Organizational Research: A Qualitative Analysis With Recommendations. Organ Res Methods. 2005;8(3):274–289. doi:10.1177/1094428105278021

26. Bernerth JB, Aguinis H. A Critical Review and Best-Practice Recommendations for Control Variable Usage. Pers Psychol. 2016;69(1):229–283. doi:10.1111/peps.12103

27. Westfall J, Yarkoni T. Statistically Controlling for Confounding Constructs Is Harder than You Think. Tran US, ed. PLOS ONE. 2016;11(3):e0152719. doi:10.1371/journal.pone.0152719

28. Wachinger C, Rieckmann A, Pölsterl S. Detect and correct bias in multi-site neuroimaging datasets. Med Image Anal. 2021;67:101879. doi:10.1016/j.media.2020.101879

29. Smith SM, Nichols TE. Statistical Challenges in “Big Data” Human Neuroimaging. Neuron. 2018;97(2):263–268. doi:10.1016/j.neuron.2017.12.018

30. Berkson J. Limitations of the Application of Fourfold Table Analysis to Hospital Data. Biom Bull. 1946;2(3):47. doi:10.2307/3002000

31. Hamdan S, Love BC, von Polier GG, et al. Confound-leakage: confound removal in machine learning leads to leakage. GigaScience. 2023;12.

32. Dinga R, Schmaal L, Penninx BWJH, Veltman DJ, Marquand AF. Controlling for Effects of Confounding Variables on Machine Learning Predictions. Bioinformatics; 2020. doi:10.1101/2020.08.17.255034

33. Berisha V, Krantsevich C, Hahn PR, et al. Digital medicine and the curse of dimensionality. Npj Digit Med. 2021;4(1):153. doi:10.1038/s41746-021-00521-5

34. Darcy AM, Louie AK, Roberts LW. Machine Learning and the Profession of Medicine. JAMA. 2016;315(6):551. doi:10.1001/jama.2015.18421

35. Jiang F, Jiang Y, Zhi H, et al. Artificial intelligence in healthcare: past, present and future. Stroke Vasc Neurol. 2017;2(4):230–243. doi:10.1136/svn-2017-000101

36. Bhinder B, Gilvary C, Madhukar NS, Elemento O. Artificial Intelligence in Cancer Research and Precision Medicine. Cancer Discov. 2021;11(4):900–915. doi:10.1158/2159-8290.CD-21-0090

37. Subramanian M, Wojtusciszyn A, Favre L, et al. Precision medicine in the era of artificial intelligence: implications in chronic disease management. J Transl Med. 2020;18(1):472. doi:10.1186/s12967-020-02658-5

38. Bzdok D, Meyer-Lindenberg A. Machine Learning for Precision Psychiatry: Opportunities and Challenges. Biol Psychiatry Cogn Neurosci Neuroimaging. 2018;3(3):223–230. doi:10.1016/j.bpsc.2017.11.007

39. Citerio G. Big Data and Artificial Intelligence for Precision Medicine in the Neuro-ICU: Bla, Bla, Bla. Neurocrit Care. 2022;37(S2):163–165. doi:10.1007/s12028-021-01427-6

40. Heinrichs B, Eickhoff SB. Your evidence? Machine learning algorithms for medical diagnosis and prediction. Hum Brain Mapp. 2020;41(6):1435–1444. doi:10.1002/hbm.24886

41. Keane PA, Topol EJ. With an eye to AI and autonomous diagnosis. Npj Digit Med. 2018;1(1):40, s41746-018-0048-y. doi:10.1038/s41746-018-0048-y

42. Topol EJ. High-performance medicine: the convergence of human and artificial intelligence. Nat Med. 2019;25(1):44–56. doi:10.1038/s41591-018-0300-7

43. Arbabshirani MR, Plis S, Sui J, Calhoun VD. Single subject prediction of brain disorders in neuroimaging: Promises and pitfalls. NeuroImage. 2017;145:137–165. doi:10.1016/j.neuroimage.2016.02.079

44. Pulini AA, Kerr WT, Loo SK, Lenartowicz A. Classification Accuracy of Neuroimaging Biomarkers in Attention-Deficit/Hyperactivity Disorder: Effects of Sample Size and Circular Analysis. Biol Psychiatry Cogn Neurosci Neuroimaging. 2019;4(2):108–120. doi:10.1016/j.bpsc.2018.06.003

45. Woo CW, Chang LJ, Lindquist MA, Wager TD. Building better biomarkers: brain models in translational neuroimaging. Nat Neurosci. 2017;20(3):365–377. doi:10.1038/nn.4478

46. Kapoor S, Narayanan A. Leakage and the Reproducibility Crisis in ML-based Science. Published online July 14, 2022. Accessed January 31, 2023. http://arxiv.org/abs/2207.07048

47. Organization WH, others. Ethics and governance of artificial intelligence for health: WHO guidance. Published online 2021.

48. Huys QJM, Maia TV, Frank MJ. Computational psychiatry as a bridge from neuroscience to clinical applications. Nat Neurosci. 2016;19(3):404–413. doi:10.1038/nn.4238

49. Weinberger DR, Radulescu E. Finding the Elusive Psychiatric “Lesion” With 21st-Century Neuroanatomy: A Note of Caution. Am J Psychiatry. 2016;173(1):27–33. doi:10.1176/appi.ajp.2015.15060753

50. O’Neil C. Weapons of Math Destruction: How Big Data Increases Inequality and Threatens Democracy. Crown; 2017.

51. Abdulkadir A, Ronneberger O, Tabrizi SJ, Klöppel S. Reduction of confounding effects with voxel-wise Gaussian process regression in structural MRI. In: 2014 International Workshop on Pattern Recognition in Neuroimaging. IEEE; 2014:1–4.

52. Dukart J, Schroeter ML, Mueller K, The Alzheimer’s Disease Neuroimaging Initiative. Age Correction in Dementia – Matching to a Healthy Brain. Valdes-Sosa PA, ed. PLoS ONE. 2011;6(7):e22193. doi:10.1371/journal.pone.0022193

53. Snoek L, Miletić S, Scholte HS. How to control for confounds in decoding analyses of neuroimaging data. NeuroImage. 2019;184:741–760. doi:10.1016/j.neuroimage.2018.09.074

54. Rao A, Monteiro JM, Ashburner J, et al. A comparison of strategies for incorporating nuisance variables into predictive neuroimaging models. In: 2015 International Workshop on Pattern Recognition in Neuroimaging.; 2015:61–64.

55. Komeyer V, Eickhoff SB, Rathkopf C, Grefkes C, Patil KR, Raimondo F. Correct deconfounding enables causal machine learning for precision medicine and beyond. Published online September 23, 2024. doi:10.1101/2024.09.20.24314055

56. Pearl J. Causal diagrams for empirical research. Biometrika. 1995;82(4):669–710.

57. Pearl J, Mackenzie D. The Book of Why: The New Science of Cause and Effect. Basic Books; 2018.

58. Sudlow C, Gallacher J, Allen N, et al. UK Biobank: An Open Access Resource for Identifying the Causes of a Wide Range of Complex Diseases of Middle and Old Age. PLOS Med. 2015;12(3):e1001779. doi:10.1371/journal.pmed.1001779

59. Bobos P, Nazari G, Lu Z, MacDermid JC. Measurement Properties of the Hand Grip Strength Assessment: A Systematic Review With Meta-analysis. Arch Phys Med Rehabil. 2020;101(3):553–565. doi:10.1016/j.apmr.2019.10.183

60. Gell M, Eickhoff SB, Omidvarnia A, et al. The Burden of Reliability: How Measurement Noise Limits Brain-Behaviour Predictions. bioRxiv. Published online February 10, 2023. doi:10.1101/2023.02.09.527898

61. Alonso AC, Ribeiro SM, Luna NMS, et al. Association between handgrip strength, balance, and knee flexion/extension strength in older adults. Sergi G, ed. PLOS ONE. 2018;13(6):e0198185. doi:10.1371/journal.pone.0198185

62. Defina S, Silva CCV, Cecil CAM, et al. Associations of Arterial Thickness, Stiffness, and Blood Pressure With Brain Morphology in Early Adolescence: A Prospective Population-Based Study. Hypertension. 2024;81(1):162–171. doi:10.1161/HYPERTENSIONAHA.123.21672

63. Kalc P, Dahnke R, Hoffstaedter F, Gaser C. Low bone mineral density is associated with gray matter volume decrease in UK Biobank. Front Aging Neurosci. 2023;15:1287304. doi:10.3389/fnagi.2023.1287304

64. Luo Y, Jiang K, He M. Association between grip strength and bone mineral density in general US population of NHANES 2013–2014. Arch Osteoporos. 2020;15(1):47. doi:10.1007/s11657-020-00719-2

65. Song J, Liu T, Zhao J, Wang S, Dang X, Wang W. Causal associations of hand grip strength with bone mineral density and fracture risk: A mendelian randomization study. Front Endocrinol. 2022;13:1020750. doi:10.3389/fendo.2022.1020750

66. Chen G, Cai Z, Taylor PA. Through the lens of causal inference: Decisions and pitfalls of covariate selection. Published online January 12, 2024. doi:10.1101/2024.01.11.575211

67. Tchernof A, Després JP, Bélanger A, et al. Reduced testosterone and adrenal C19 steroid levels in obese men. Metabolism. 1995;44(4):513–519. doi:10.1016/0026-0495(95)90060-8

68. He et al. -2022 - Meta-matching as a simple framework to translate p.pdf.

