## Supplementary Material for "Confounder control in biomedicine necessitates conceptual considerations beyond statistical evaluations"

*Contents:*

Methods
Supplementary Table S1
Code availability
Supplementary figures
Supplementary references

### 1. Methods

#### 1.1. Data and pre-processing

For the example predictions we used data of the 1st scanning session (ses-2) of the UK Biobank[1], recorded at three different sites in the UK (Cheadle, Reading, Newcastle). The exact protocol and acquisition parameters of both the structural imaging as well as the rs-fMRI can be found in Miller et al. (2016)[2]. The initital structural and functional pre-processing was carried out by pipelines developed and run by the UKB[3].

For the grey matter volume (GMV) features 41,180 T1-weighted pre-processed images were retrieved from UKB and converted into a DataLad[4] dataset for provenance tracking with subsequent computations of voxel-based morphometry (CAT 12.7 (default settings); MNI152 space; 1.5mm isotropic)[5]. We extracted the parcel-wise GMV as the winsorized mean (limits 10%) of the voxel-wise values per parcel using the cortical Schaefer et al. (2018)[6] atlas (1000 ROIs), subcortical Tian et al. (2020)[7] (S4 3T) and cerebellar Diedrichsen et al. (2009)[8] (SUIT space) atlas. To retrieve the functional connectivity (FC) features, 5000 subjects of the UKB pre-processed rs-fMRI datasets were normalized to MNI space using FSL. After denoising, each subject's time course was parcellated with the Schaefer et al. (2018) cortical atlas (400 ROIs) by averaging across all voxels of each parcel. The FC was eventually calculated as the Pearson correlations of the parcel-wise time-series.

All non-imaging variables, including the exemplarily target hand grip strength (HGS) and the investigated example confounders were obtained directly from the UK Biobank[13]. We chose HGS as a robust, objective and reliable target[14–17] to avoid further conceptual problems oftentimes coming along with more latent variables as targets, such as intelligence or executive functioning measures[18]. Healthy subjects were (rather conservatively) defined by excluding the ICD-10 criteria chapters F, G and I60 to I69, which excludes subjects with a history of mental and behavioural disorders, diseases of the nervous system or with a cerebrovascular disease. All NaN values and outliers larger than the 4th standard deviation were removed from the non-imaging data. Additionally, the HGS was averaged over left and right hand and there was a check for balance of sex distributions in the HGS.

#### 1.2. Modelling Setup

10% of the data were set apart to be used as a locked test set for a related project and left untouched for this project. The remaining 90% of the data were split into a training (0.8) and test (0.2) set. The learning algorithms were then fitted on the training set by using a stratified 5-fold cross-validation (CV) scheme with one repetition. Hyperparameters were tuned within a nested CV, i.e. to each fold of the outer CV a shuffled 5-fold CV scheme was applied to find the best hyperparameters. This was achieved through a grid search with RMSE (root mean squared error) error metric to identify the best hyperparameter. The outer CV on the training set served to control for the fitting behaviour (e.g. overfitting) of the model and to control for the generalization error. A final estimator, retrained on the entire training set (using RMSE) was eventually used to make the predictions on the initially held-out test set. These predictions were used to report and visualize the predictive performances in Figure 1 of the manuscript. All applied splits were stratified for binned age, binned HGS (2 bins) and sex (as either defined in the NHS central registry or self-reported). Within the CV scheme, continuous features and confounders were z-scored (mean of zero and unit variance) and categorical confounders (sex) were one-hot-encoded. Z-scoring the confounds didn't make a difference in the predictions. Outer CV performance was evaluated using RMSE, mean absolute error (MAE), coefficient of determination ($R^2$), Pearson's r and Spearman's r. The confound removal was applied within the CV to avoid data leakage. Therein, for each feature, a linear regression was fit using the confounds as independent variables and the features as dependent variables. The new, confound-free features were calculated as the residuals of the fitted linear regression (original features minus predicted/fitted features).

#### 1.3. Algorithms and sample sizes

For the ridge regression used for the "vanilla model" and the ones showing the feature and confounder impact we use scikit-learn's[20] RidgeCV implementation that comes with an in-built and optimized nested CV. We tuned the best hyperparameter alpha, i.e. the amount of regularization, within a grid of [10, 100, 1e3, 1e4, 1e5, 1e6] and identified the best hyperparameter through RMSE scoring. As comparison algorithm for the “algorithm impact”-model a grid-search CV was used to find the best hyperparameters for a support vector regression (SVR) with the non-linear radial basis function (RBF) kernel (C in [0.5, 0.1, 0.3]; gamma = ‘scale’; epsilon in [.1, .5, .6]).

For the GMV features with the linear ridge regression and the non-linear SVR(RBF) algorithm we used the data of N=27 846 subjects ($N_{train}$=22 276, $N_{test}$=5 570) (Table S1). For the sex and age adjusted ridge regression data of N=23 242 ($N_{train}$=18 593, $N_{test}$=4 649) subjects was used and the FC model was trained and tested with N=4 172 ($N_{train}$=3 337, $N_{test}$=835) subjects (Table 1).

**Table S1.** Overview of sample sizes.

| | $N_{train}$ | $N_{test}$ (hold-out) |
|---|---|---|
| **GMV**<br>**Ridge Regression (linear)**<br>○ ***Vanilla***<br>○ **Sex & Age adjusted** | <br><br>22,276<br>18,593 | <br><br>5,570<br>4,649 |
| **GMV**<br>**SVR(RBF) (non-linear)**<br>**No confound removal** | 22,276 | 5,570 |
| **FC**<br>**Ridge Regression (linear)**<br>**No confound removal** | 3,337 | 835 |

##### 1.4. Statistics for Confound Continuum

To investigate the statistical confound-feature and confound-target relationship, we broadly inspected non-imaging variables in the UKB and narrowed them down to 130 problem-related variables, sorted into 12 higher-level categories (colour coding in Figure 2A of the manuscript). Each variable was then (independently) correlated with parcel-wise GMV features and HGS (Figure S1), using Pearson’s r for continuous confounds, Spearman rank correlation for ordinate variables and a point biserial correlation for binary variables. For the variables sex, age, scan-site and scan-time we additionally investigated the distribution of the parcel-wise correlation coefficients and checked their anatomical positions. The main manuscript shows only correlations with the target, with reduced annotations, while Figure S1 visualizes the correlations of the 130 selected variables with both, parcellated GMV features (Figure S1B) and the target HGS (Figure 1S A), including detailed annotations. Grey bars in Figure S1 are only for improved readability of the graph and do not imply statistical meaning.

##### 1.5. Code availability

All corresponding codes together with additional information on code execution and replication can be found on GitHub under https://github.com/verakye/ConfoundContinuum.

#### 2. Supplementary figures

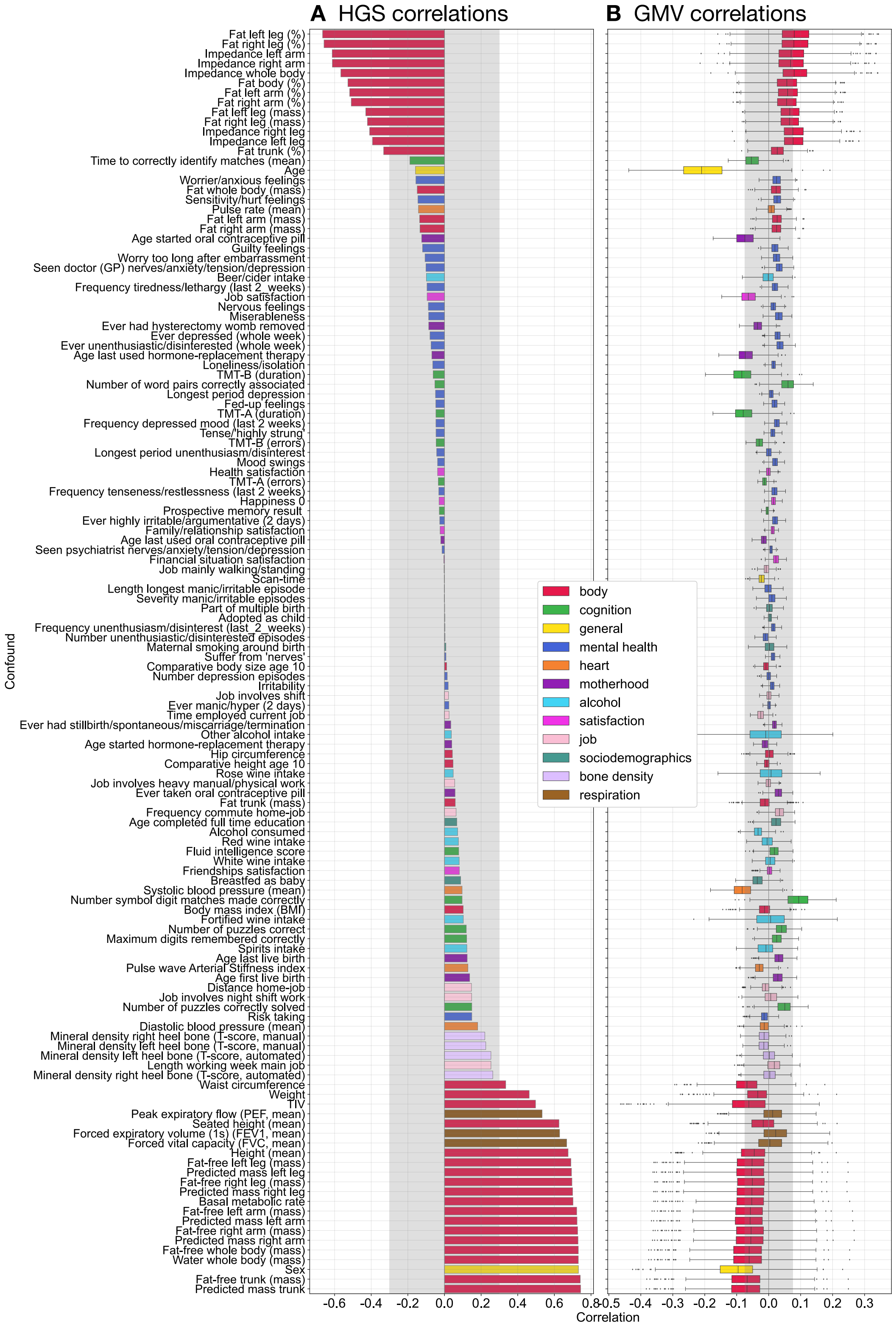


**Figure S1.** Correlations of 130 summary behavioural variables with the exemplary target HGS (**A**) and the 1088 parcellated GMV features (**B**) that could potentially be considered as confounding variables. The variables were sorted into 12 higher-level categories. Boxplots in B) indicate median (IQR) correlation over GMV parcels. Correlations refer to Pearson's r for continuous confounds, Spearman correlations for ordinal variables and point-biserial correlation coefficients for binary variables. Grey areas indicate absolute correlations smaller than |r|=0.3 for HGS (A) and |r|=0.075 for GMV (B) for better readability and visualization of correlation strengths.

#### 3. References

1. Sudlow C, Gallacher J, Allen N, et al. UK Biobank: An Open Access Resource for Identifying the Causes of a Wide Range of Complex Diseases of Middle and Old Age. *PLOS Med*. 2015;12(3):e1001779. doi:10.1371/journal.pmed.1001779

2. Miller KL, Alfaro-Almagro F, Bangerter NK, et al. Multimodal population brain imaging in the UK Biobank prospective epidemiological study. *Nat Neurosci*. 2016;19(11):1523-1536. doi:10.1038/nn.4393

3. Alfaro-Almagro F, Jenkinson M, Bangerter NK, et al. Image processing and Quality Control for the first 10,000 brain imaging datasets from UK Biobank. *NeuroImage*. 2018;166:400-424. doi:10.1016/j.neuroimage.2017.10.034

4. Halchenko YO, Meyer K, Poldrack B, et al. DataLad: distributed system for joint management of code, data, and their relationship. *J Open Source Softw*. 2021;6(63):3262. doi:10.21105/joss.03262

5. Wagner AS, Waite LK, Wierzba M, et al. FAIRly big: A framework for computationally reproducible processing of large-scale data. :25.

6. Schaefer A, Kong R, Gordon EM, et al. Local-Global Parcellation of the Human Cerebral Cortex from Intrinsic Functional Connectivity MRI. *Cereb Cortex*. 2018;28(9):3095-3114. doi:10.1093/cercor/bhx179

7. Tian Y, Margulies DS, Breakspear M, Zalesky A. Topographic organization of the human subcortex unveiled with functional connectivity gradients. *Nat Neurosci*. 2020;23(11):1421-1432. doi:10.1038/s41593-020-00711-6

8. Diedrichsen J, Balsters JH, Flavell J, Cussans E, Ramnani N. A probabilistic MR atlas of the human cerebellum. *NeuroImage*. 2009;46(1):39-46. doi:10.1016/j.neuroimage.2009.01.045

9. Fischl B. FreeSurfer. *NeuroImage*. 2012;62(2):774-781. doi:10.1016/j.neuroimage.2012.01.021

10. FreeSurfer. FreeSurfer. Accessed February 16, 2023. https://surfer.nmr.mgh.harvard.edu

11. Dale AM, Fischl B, Sereno MI. Cortical Surface-Based Analysis.

12. Desikan RS, Ségonne F, Fischl B, et al. An automated labeling system for subdividing the human cerebral cortex on MRI scans into gyral based regions of interest. *NeuroImage*. 2006;31(3):968-980. doi:10.1016/j.neuroimage.2006.01.021

13. Brandes N, Linial N, Linial M. PWAS: proteome-wide association study—linking genes and phenotypes by functional variation in proteins. *Genome Biol*. 2020;21(1):173. doi:10.1186/s13059-020-02089-x

14. Alonso AC, Ribeiro SM, Luna NMS, et al. Association between handgrip strength, balance, and knee flexion/extension strength in older adults. Sergi G, ed. *PLOS ONE*. 2018;13(6):e0198185. doi:10.1371/journal.pone.0198185

15. Bobos P, Nazari G, Lu Z, MacDermid JC. Measurement Properties of the Hand Grip Strength Assessment: A Systematic Review With Meta-analysis. *Arch Phys Med Rehabil*. 2020;101(3):553-565. doi:10.1016/j.apmr.2019.10.183

16. Bohannon RW, Schaubert KL. Test–retest reliability of grip-strength measures obtained over a 12-week interval from community-dwelling elders. *J Hand Ther*. 2005;18(4):426-428.

17. Reuter SE, Massy-Westropp N, Evans AM. Reliability and validity of indices of hand-grip strength and endurance: EVALUATION OF GRIP STRENGTH AND ENDURANCE. *Aust Occup Ther J*. 2011;58(2):82-87. doi:10.1111/j.1440-1630.2010.00888.x

18. Gell M, Eickhoff SB, Omidvarnia A, et al. *The Burden of Reliability: How Measurement Noise Limits Brain-Behaviour Predictions*. Neuroscience; 2023. doi:10.1101/2023.02.09.527898

19. R: Fast Heuristics For The Estimation Of the C Constant Of A... Accessed December 9, 2022. https://search.r-project.org/CRAN/refmans/LiblineaR/html/heuristicC.html

20. Pedregosa F, Varoquaux G, Gramfort A, et al. Scikit-learn: Machine Learning in Python. *Mach Learn PYTHON*.:6.

21. Fan RE, Chang KW, Hsieh CJ, Wang XR, Lin CJ. LIBLINEAR: A Library for Large Linear Classification. :33.

22. Virtanen P, Gommers R, Oliphant TE, et al. SciPy 1.0: Fundamental algorithms for scientific computing in python. *Nat Methods*. 2020;17:261-272. doi:10.1038/s41592-019-0686-2

23. Vallat R. Pingouin: statistics in Python. *J Open Source Softw*. 2018;3(31):1026. doi:10.21105/joss.01026
